# Cross-sectional Surveys: Impact of Kidney-CAP availability on health provider and patient decisions related to vascular access, dialysis modality and kidney transplantation

**DOI:** 10.64898/2026.03.09.26347976

**Authors:** Adam Forster, Faisal Rehman, Louise Moist, Rachel Holden, Benjamin Thomson

## Abstract

**Introduction:** Catastrophic bleeding can be fatal in patients on hemodialysis using Arteriovenous (AV) fistulas or grafts. Campaigns, such as the UK “Put a Lid On It” and the Australia “Stop the Bleed” have recommended use of bleeding cessation devices, but evidence for their use remains limited. Recent creation of the bleeding cessation device “Kidney-CAP” mandated further study. The objective of this study was to determine how the Kidney-CAP modified decisions related to vascular access, dialysis modality, and kidney transplantation.

**Methods:** Cross-sectional surveys were administered at a Canadian academic nephrology program, to health care providers (HCP) managing patients with chronic kidney disease (CKD), to patients on hemodialysis (CKD-HD), and to patients with CKD but not on dialysis (CKD-Clinic). Two tailed, one sample sign test was used to determine if the median response to Likert scale questions differed from “no effect” response, to a p-value of < 0.05.

**Results:** Survey respondents included 18 HCP, 23 CKD-HD and 30 CKD-Clinic patients. Having a Kidney-CAP increased CKD-Clinic patients’ desire to undergo AVF or AVG creation (p=0.020). Having a Kidney-CAP had no impact on CKD-HD patients’ deside to undergo AVF creation, or to pursue hemodialysis at home, but increased desire to undergo kidney transplantation (p=0.031). Availability of the Kidney-CAP had no impact on HCP recommendations related to AVF creation or kidney transplantation, but increased HCP recommendations for home hemodialysis in ESKD patients (p=0.039 for each).

**Conclusion:** This is the first study to assess the perceived benefit of a bleeding cessation device, with a focus on clinical decision making related to vascular access, kidney transplantation, and dialysis modality. The Kidney-CAP is associated with increased patient uptake of kidney transplantation and creation of AVF. Further study is required to delineate patient decisions within demographic subgroups such as previous kidney transplant, or anticoagulation status.

## INTRODUCTION

Hemodialysis patients with an arteriovenous fistula (AVF) or arteriovenous graft (AVG) are at increased risk of hemorrhage because of coagulopathy of renal disease, and hemodialysis circuit anticoagulation.(1, 2) There are best practice guidelines for management of prolonged bleeding after needle removal from AVF/AVG, when a patient is in the hemodialysis facility.(3, 4, 5, 6) However, these guidelines do not address AVF/AVG bleeding outside the hemodialysis facility, other than stating that this is a surgical emergency.(5, 6) AVF/AVG Bleeding outside the hemodialysis unit can be catastrophic,(7, 8) and thus optimizing management strategies is important. A recently published scoping review identified a paucity of research on methods to manage AVF/AVG bleeding outside the hemodialysis unit, and called for more research on the topic.(9)

The UK “Put a Lid On It!”(10) and the Australia “Stop the Bleed”(11) campaigns provided a device to hemodialysis patients, to manage AVF/AVG bleeding outside the hospital. While the campaigns reported widespread patient acceptance of the devices, whether they impact clinical outcomes is uncertain. Fear of AVF/AVG cannulation and bleeding are common with hemodialysis patients, and may alter decisions related to vascular access, dialysis modality, and kidney transplantation.(12, 13, 14, 15) However, it is unknown if availability of a device designed to stop AVF/AVG bleeding outside of the dialysis facility modifies these patient decisions. Similarly, it’s unknown if nephrology health providers modify recommendations to patients, based on the device availability.

Starting in 2022, Glia organization (https://glia.org/pages/glia-kidney-cap) designed a 3D-printed device (Kidney-CAP) to stop AVF/AVG bleeding outside the dialysis unit. The Kidney-CAP design was informed by input from nephrology health providers, patients, engineers, and 3D-printing experts. Glia has secured approval from Health Canada and the Food and Drug Administration to provide the Kidney-CAP to patients. However, further study is needed to justify the Kidney-CAP’s broader clinical adoption.

This study had two objectives. First, it examined how the availability of the Kidney-CAP influenced decisions related to vascular access, dialysis modality, and kidney transplantation, in patients with advanced chronic kidney disease (CKD) on hemodialysis (CKD-HD) or being followed in a multidisciplinary advanced CKD- non dialysis (CKD-Clinic). Second, we assessed how Kidney-CAP availability modified nephrology HCP recommendations on vascular access, dialysis modality, and kidney transplantation for patients with CKD

## MATERIALS and METHODS

Three cross-sectional surveys were designed to assess how Kidney-CAP availability modified patient decisions and health care provider (HCP) recommendations related to vascular access, dialysis modality, and kidney transplantation. Each survey was preceded by an information page that included a photograph and instructions on Kidney-CAP use (Appendix 1). Likert scale (significantly decrease, decrease, no effect, increase or significantly increase) was used to evaluate effect on the likelihood of recommending or being comfortable with kidney transplantation, creation of AVF or home hemodialysis. Likert scale surveys have been used previously to report nephrology patient(16, 17, 18, 19) and health care provider(20, 21) preferences. The study was reported according to the Consensus-Based Checklist for Reporting of Survey Studies (CROSS).(22) Survey recruitement period was between January 23 (2024) and October 8 (2025). Written consent was obtained prior to beginning the survey, and was documented electronically with the survey answers.

That the Kidney-CAP avaialability could impact decisions related to kidney transplantation may not be intuitive. However, patient decisions regarding kidney transplantation are complex, and modified by both the available dialysis modality choices, and also patient experience while on dialysis.(23) Given an impact of the Kidney-CAP on transplant related choices could not be excluded, the surveys included assessment of this domain.

### Survey 1- Health Care Providers (HCP)

Cross-sectional survey 1 (HCP) provided scenarios of patients that differed only by stage of CKD or vascular access (Case 1: CKD stage G5A2 with no vascular access plan; Case 2: ESKD-HD using central venous catheter; Case 3: ESKD-HD with well-functioning AVF; Case 4: ESKD-HD with prolonged bleeding from AVF). Survey questions evaluated the likelihood of HCP recommending home hemodialysis, kidney transplantation, or (in cases 1 and 2) AVF creation. Other questions evaluated when and who to introduce patients to the Kidney-CAP, demographic factors of survey respondents, and level of concern related to AVF/AVG bleeds outside the dialysis unit.

### Sampling and recruitment

Health Care providers (HCP) at London Health Sciences Centre and in the Division of Nephrology at the University of Western Ontario (London, Ontario, Canada) include nephrologists, vascular access coordinators, hemodialysis nurse practitioners, nephrology clinic or hemodialysis nurses. Survey 1 (HCP) was emailed to all department members with a link to the survey provided on REDCap (Nashville, USA) survey software. Two reminder emails were sent to complete the survey, 2 and 4 weeks after the initial email, to facilitate a higher response rate.

### Survey 2: Chronic Kidney Disease patients on hemodialysis (CKD-HD)

Cross-sectional survey 2 (CKD-HD) first identified hemodialysis vintage and frequency, exposure to antiplatelets or anticoagulant medications, current vascular access, prior kidney transplantation, and duration of nephrology follow-up. When patients had a history of AVF or AVG, a bleeding history was established. The impact of Kidney-CAP availability on patent decisions related to kidney transplantation, home hemodialysis, or (when patient had central venous catheter) creation of AVF or AVG was evaluated. Additional demographic factors were also established.

### Sampling and recruitment

Research coordinators approached patients in the hemodialysis unit (CKD-HD) at London Health Sciences Centre with an iPad, on two consecutive hemodialysis shifts, on two consecutive days. The iPad had the survey available through REDCap (Nashville, USA) survey software, and the research coordinator remained available to study participants if challenges with literacy or vision were identified.

### Survey 3: Chronic Kidney Disease patients in multidisciplinary kidney disease clinic (CKD-Clinic)

Cross-sectional survey 3 (CKD-Clinic) first evaluated exposure to antiplatelet or anticoagulant medications, duration of nephrology follow-up, history of kidney transplantation, and other demographic factors (eg. Age, sex, race, marital status). The impact of Kidney-CAP availability on decisions related to vascular access, home hemodialysis and kidney transplantation was then assessed.

### Sampling and recruitment

Research coordinators approached patients in the multidisciplinary kidney disease clinic (CKD-Clinic) at London Health Sciences Centre with an iPad, on two separate days. To attend the multidisciplinary kidney disease clinic (CKD-Clinic), patients needed to have at least 10.0% chance by Kidney Failure Risk Equation of requiring renal replacement therapy within 2 years,(24) as per provincial guidelines.(25) The iPad had the survey available through REDCap (Nashville, USA) survey software, and the research coordinator remained available to study participants if challenges with literacy or vision were identified.

A copy of each survey is available by request to the study’s corresponding author.

### Survey validation

The research goals and cross-sectional surveys were first reviewed by 3 nephrologists (Survey 1) and 2 patients (Surveys 2 and 3) to assure face validity. The surveys were then reviewed with 3 researchers with experience in survey research methodology. This second review was to enhance criterion and construct validity.

Duplicate entries by a single respondent are prevented in REDCap by preventing a single IP address from duplicate entries, and also by asking respondents to complete the survey only once.

### Data collection and analysis

Data collection was automated in REDCap (Nashville, USA), then data transferred for analysis in StataMP 18.0 (College Station, Texas USA).

### Missing data

Patients were permitted not to answer a question if they chose. Missing data was managed by the listwise deletion method.

### Likert Scale answers from Surveys 1,2,3

The two-tailed, one sample sign test was used to determine if the median response was significantly different from neutral “no effect,” “neither easy nor difficult” or “the right timing” response, with p<0.05 for statistical significance.

### Ethics

This study was approved by the Health Sciences Research Ethics Board at Western University (REB#121690). Survey respondents remained anonymous throughout survey and study completion, with confidentiality maintained.

## RESULTS

### Survey 1 (Health Care Providers)

There were 18 nephrology health care provider respondents, which included nephrology clinic and hemodialysis nurses (n=9, 50.0%; response rate 37.5%), nephrology nurse practitioners (n=4, 22.2%; response rate 66.7%), nephrologists (n=4, 22.2%; response rate 23.5%) and vascular access coordinator (n=1, 5.6%; response rate 50.0%). The median experience working in nephrology was 12 years (range 4 to 15 years).

Availability of Kidney-CAP had no impact on the likelihood of health care providers recommending patient’s undergo AVF creation, in either of the two cases (CKD stage G5A2, or ESKD on hemodialysis for one month using CVC) (Figure 1).

**Figure 1:**
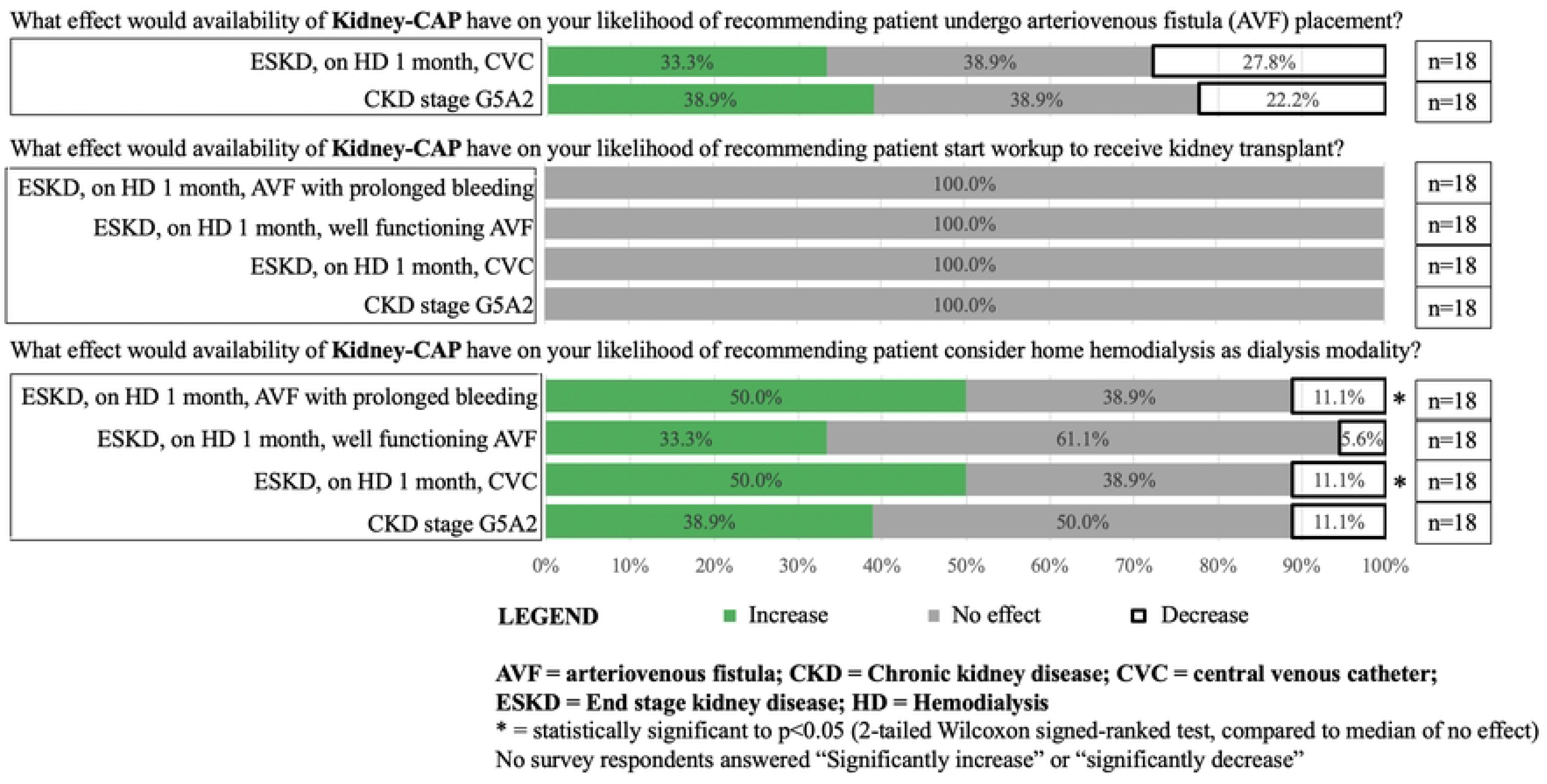
Impact of Availability of Kidney-CAP on Nephrology Clinical Care Provider Recommendations. **LEGEND** 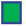Increase 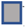No effect 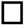Decrease AVF = arteriovenous fistula; CKD = Chronic kidney disease; CVC = central venous catheter; ESKD = End stage kidney disease; HD = Hemodialysis * = statistically significant to p<0.05 (2-tailed Wilcoxon signed-ranked test, compared to median of no effect) No survey respondents answered “Significantly increase” or “significantly decrease”

The availability of Kidney-CAP did not affect the likelihood that HCP would recommend initiating a kidney transplant work up , in any of the four cases (Figure 1).

The availability of the Kidney-CAP increased the likelihood that HCPs would recommend home hemodialysis for patients with ESKD who have been on HD for 1 month using central venous catheter (CVC) (p=0.039) and for patients on hemodialysis for one month using an AVF who were experiencing prolonged bleeding (p=0.039) (Figure 1)

Most respondents (13/18) felt that providing education on the Kidney-CAP to patients with CKD stage G5A2 was either slightly or very early (Figure 2). In contrast, most respondents indicated that providing education on the Kidney-CAP was slightly or very late for most respondents (14/18) when patients had ESKD, on hemodialysis for 1 month with an AVF with prolonged bleeding. A median response of “the right timing” was found only for patients with ESKD, on hemodialysis for one month using a CVC.

**Figure 2:**
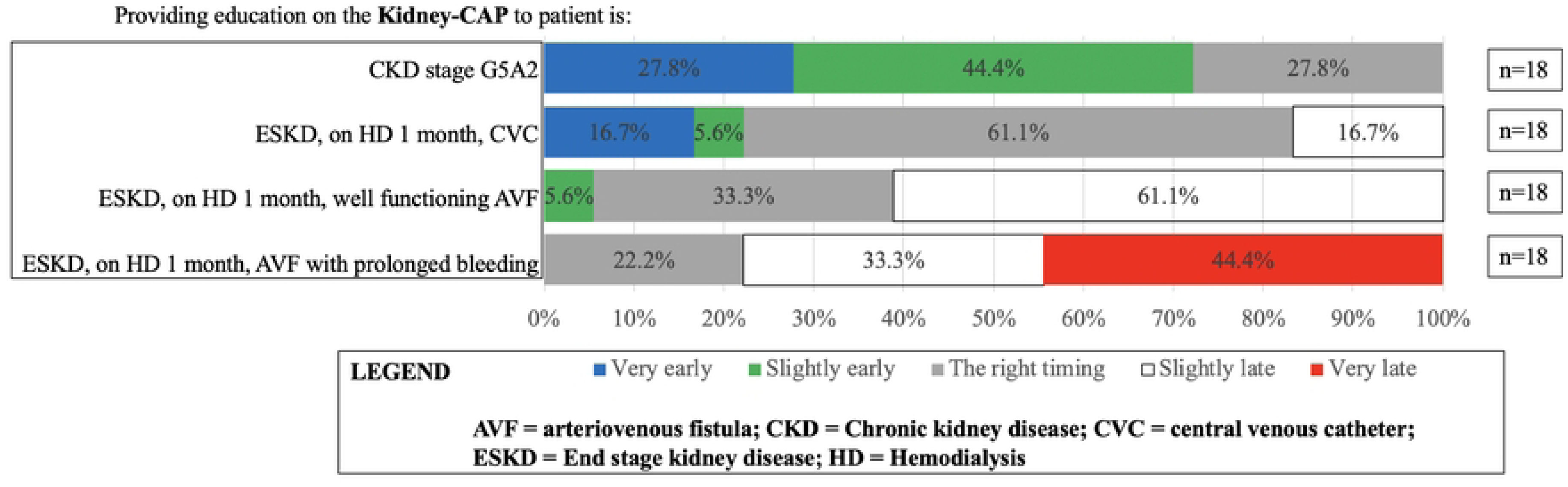
Nephrology Provider Recommendations for Timing for Kidney-CAP education. **LEGEND** 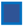Very early 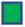Slightly early 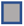The right timing 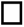Slightly late 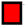Very late **AVF = arteriovenous fistula; CKD = Chronic kidney disease; CVC = central venous catheter; ESKD = End stage kidney disease; HD = Hemodialysis**

### Survey 2 (CKD-Hemodialysis)

There were 23 respondents (Table 1), from 268 patients approached. Most respondents were Caucasian (69.6%, n=16) and female (60.9%), with mean age 59 years. The top three highest attained education levels were high school diploma (30.4%, n=7), college diploma (26.1%, n=6) and bachelor’s degree (17.4%, n=4). The median number of people per household was 2, with half the respondents having been followed by nephrology for 9 or more years. Most respondents were single (26.1%, n=6), married (26.1%, n=6) or widowed (26.1%, n=6). Warfarin (8.6%, n=2) and DOAC (direct oral anticoagulant)(17.4%, n=4) use was noted in a minority of respondents. Several respondents had undergone prior kidney transplant (18.2%, n=4).

**Table 1:** Demographic factors for Chronic Kidney Disease (CKD-Clinic) and Hemodialysis (CKD-HD) patients.

| Parameter | Chronic Kidney Disease (n=30) | Hemodialysis (n=23) |
| --- | --- | --- |
| Age | 67 (62-72) | 59 (44-66) |
| Female | 14 (46.7%) | 14 (60.9%) |
| Race |  |  |
| *Black* | 4 (13.3%) | 2 (8.7%) |
| *Indigenous or First Nations* | 2 (6.7%) | 2 (8.7%) |
| *White or Caucasian* | 21 (70%) | 16 (69.6%) |
| *Hispanic or Latino* | 0 (0%) | 1 (4.3%) |
| *Asian* | 3 (10%) | 1 (4.3%) |
| *Prefer not to answer* | 0 | 1 (4.3%) |
| Education Level |  |  |
| *No certificate, diploma, or degree* | 0 (0.0%) | 1 (4.3%) |
| *High school diploma* | 14 (46.7%) | 7 (30.4%) |
| *Apprenticeship* | 1 (3.3%) | 1 (4.3%) |
| *College diploma* | 3 (10%) | 6 (26.1%) |
| *University below Bachelor's* | 2 (6.7%) | 1 (4.3%) |
| *Bachelor's degree* | 9 (30%) | 4 (17.4%) |
| *Graduate degree* | 1 (3.3%) | 1 (4.3%) |
| *Prefer not to answer* | 0 (0.0%) | 2 (8.7%) |
| Number of people in household | 2 (2-2) | 2 (1-2.5) |
| Median household income bracket | 40445 – 49106 | 49107 – 92454 |
| Years followed by nephrology | 5 (3.25-6) | 9 (4-11) |
| Marital status |  |  |
| *Single* | 1 (3.3%) | 6 (26.1%) |
| *In a relationship, not living together* | 1 (3.3%) | 1 (4.3%) |
| *Common law partner* | 5 (16.7%) | 1 (4.3%) |
| *Marriage* | 20 (66.7%) | 6 (26.1%) |
| *Separated or divorced* | 1 (3.3%) | 2 (8.6%) |
| *Widowed* | 2 (6.7%) | 6 (26.1%) |
| *Prefer not to answer* | 0 (0.0%) | 1 (4.3%) |
| Blood thinner use |  |  |
| *Direct oral anticoagulant* | 6 (20%) | 4 (17.4%) |
| *Warfarin* | 0 (0.0%) | 2 (8.6%) |
| *Aspirin* | 7 (23.3%) | 2 (8.6%) |
| *Other antiplatelet* | 0 (0.0%) | 1 (4.3%) |
| Previous kidney transplant | 1 (3.3%) | 4 (18.2%) |

Having knowledge of the Kidney-CAP had no impact on CKD-Hemodialysis patients’ desire to undergo creation of a hemodialysis AVF or AVG, or to pursue hemodialysis at home (Figure 3). However, having a Kidney-CAP increased desire to undergo kidney transplantation (p=0.031).

**Figure 3:**
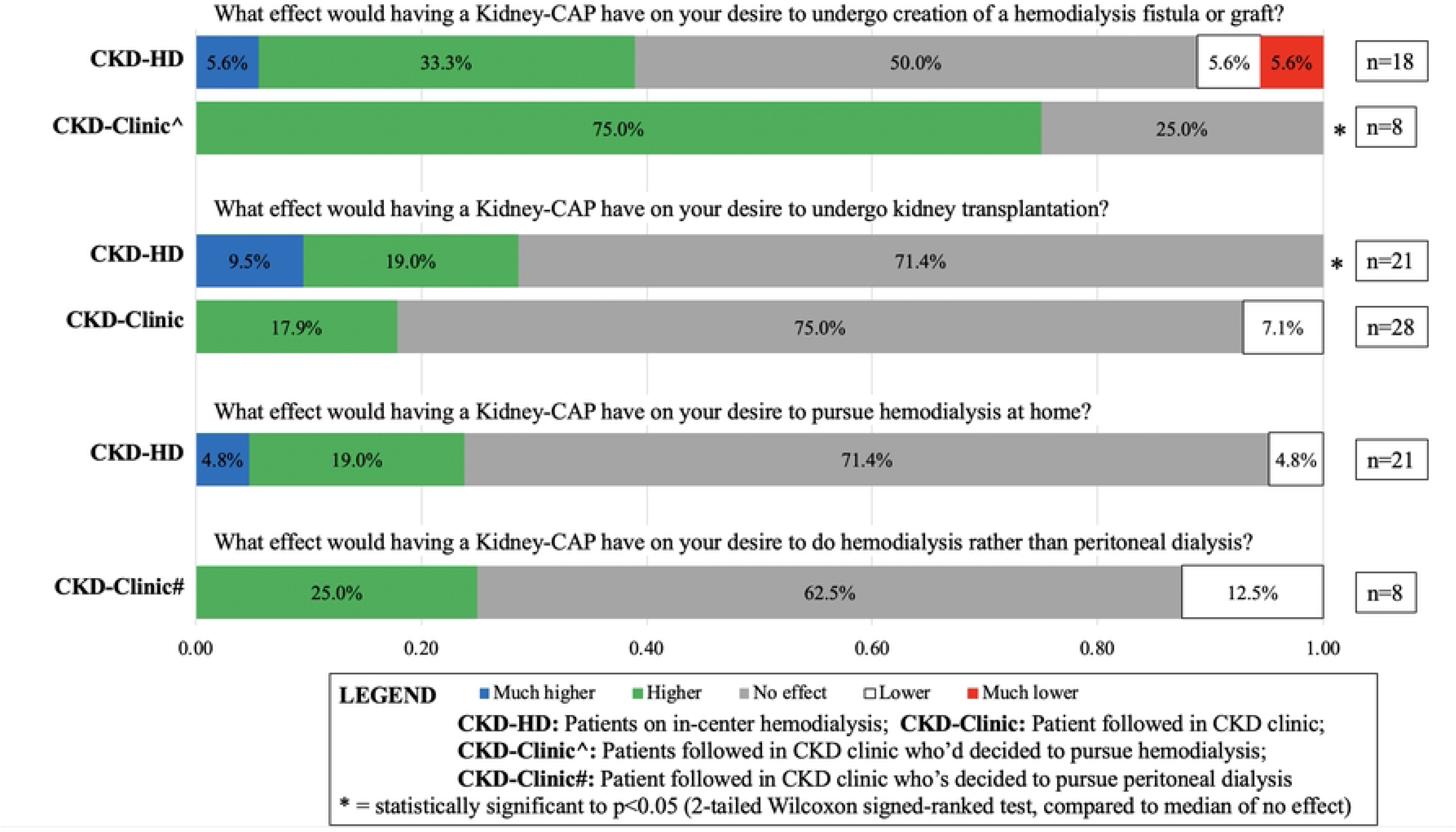
Impact of Availability of Kidney-CAP on Survey Respondents’ Clinical Decisions. **LEGEND** 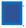Much higher 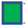Higher 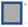No effect 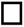Lower 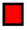Much lower **CKD-HD:** Patients on in-center hemodialysis; CKD-Clinic: Patient followed in CKD clinic; **CKD-Clinic^:** Patients followed in CKD clinic who’d decided to pursue hemodialysis; **CKD-Clinic#:** Patient followed in CKD clinic who’s decided to pursue peritoneal dialysis **\* =** statistically significant to p<0.05 (2-tailed Wilcoxon signed-ranked test, compared to median of no effect)

Most patients felt using the Kidney-CAP to stop bleeding from a hemodialysis fistula or graft would be very easy or easy (p=0.001) (Figure 4).

**Figure 4:**
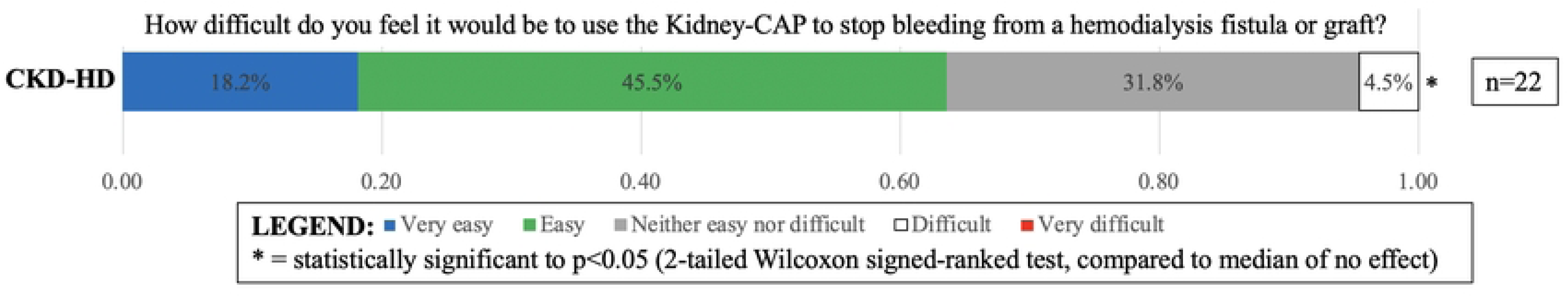
Ease of use of Kidney-CAP LEGEND. 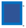Very easy 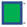Easy 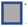Neither easy nor difficult 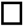Difficult 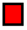 Very difficult * = statistically significant to p<0.05 (2-tailed Wilcoxon signed-ranked test, compared to median of no effect)

### Survey 3 (CKD-Clinic)

There were 30 respondents (Table 1), from 91 patients approached. Most respondents were Caucasian (70.0%, n=21) and male (53.3%, n=16). The top three highest attained education levels were high school diploma (46.7%, n=14), Bachelor’s degree (30.0%, n=9) and college diploma (10.0%, n=3). The median number of people per household was 2, with half the respondents having been followed by nephrology for 5 or more years. Most respondents were married (66.7%, n=20), common law partner (16.7%, n=5) or widowed (6.7%, n=2). None of the patients used warfarin, but DOAC (20.0%, n=6) and Aspirin (23.3%, n=7) use was common. Only 1 respondent (3.3%) had undergone previous kidney transplantation.

Having knowledge of the Kidney-CAP had no impact on CKD-Clinic patients’s desire to undergo kidney transplantation, or (in a subset who had chosen to pursue peritoneal dialysis) to change their decision to hemodialysis (Figure 3). However, Kidney-CAP awareness increased desire to undergo creation of a hemodialysis AVF or AVG (p=0.020).

## DISCUSSION

There has been increased use of bleeding cessation devices for hemodialysis patients with AVF/AVG,(10, 11) but evidence supporting their use has been lacking. When outside the hemodialysis unit, bleeding from AVF/AVG can be fatal,(7, 8) and thus evidence is critically needed. This is the first study that evaluates the impact of the availability of a bleeding cessation device, called “Kidney-CAP,” on the decisions that health care providers and patients make along their chronic kidney disease journey.

Kidney-CAP availability had no impact on health care providers’ likelihood of recommending kidney transplantation, in any of 4 patient cases. However, hemodialysis patients with a Kidney-CAP were more likely to want to undergo kidney transplantation. This study did not evaluate the reasons for this conclusion; however, chronic hemodialysis patients believe that kidney transplantation improves quality of life and extends life.(26) Therefore, having a Kidney-CAP may remind hemodialysis patients of the risks of potentially fatal bleeding, incentivizing kidney transplantation. On the other hand, decisions related to transplant were not affected by the Kidney-CAP in CKD patients not on dialysis. Patients may need to experience bleeding from an AVF or AVG, in the hemodialysis unit, before it has a meaningful impact on decision making regarding kidney transplantation. Further study is required.

Kidney-CAP availability had no impact on health care providers’ likelihood of recommending AVF creation, but increased the desire to undergo AVF creation in patients followed in CKD clinic (not yet on dialysis) whose chosen dialysis modality was hemodialysis. Health care providers and patients have different priorities in the vascular access decision making process, with patients and their families having significant fears of aneurysms or bleeding following AVF use(27, 28, 29); Kidney-CAP availability may help to alleviate these patient fears.

It is an important finding that Kidney-CAP availability increased patients’ choosing to undergo AVF creation if that patient had already chosen to pursue hemodialysis. A systematic review failed to identify evidence-based interventions to increase use of AVF/AVG in incident hemodialysis patients.(30) However, a patient’s decision to undergo AVF creation is modified by fear of cannulation and bleeding.(31, 32) Patients who refuse to undergo AVF creation have longer dialysis vintage, (33) and are thus more likely to have seen or heard about a bleeding complication from an AVF. Kidney-CAP may address these fears and make patients more open to undergoing AVF creation.

The willingness to participate in the cross-sectional surveys was low in both CKD-HD (23/268) and CKD-Clinic (30/91) patients. Willingness to participate in clinical trials is influenced not only by patient demographic factors but also study factors. Hemodialysis patients willing to participate tend to be younger, nonsmokers, more likely to be recently hospitalized, less likely to be on a kidney transplant waiting list, and less likely to have either reactive airway disease or coronary artery disease.(34) Data were not collected for non-participants of our study, so whether demographic factors impacted participation is unknown. On the other hand, patients were not promised to be receive a Kidney-CAP at the end of the study; This could have been perceived similarly to placebo use, which decreases participation.(35, 36) The inclusion of the Kidney-CAP device and the 1-page training sheet may have been interpreted as overly complex, again decreasing study participation.(37)

This study evaluated when to introduce the Kidney-CAP in the pathway of chronic kidney disease care. Health care providers agreed that it would be too early to introduce the Kidney-CAP to a non-dialysis CKD patient who’d not yet made a dialysis modality decision. They also felt that it was too late to introduce it to a hemodialysis patient with an AVF with prolonged post-cannulation bleeding, but also to a hemodialysis patient with a well-functioning AVF. The best time to introduce the Kidney-CAP is likely when a patient with chronic kidney disease has made a choice to pursue hemodialysis, and is evaluating vascular access options. This would also yield the benefit of the Kidney-CAP device in patients with CKD clinic who’d chosen to pursue hemodialysis; in this group of patients having the Kidney-CAP increased the desire to undergo AVF or AVG creation. In that context, the Kidney-CAP may address patient concerns of bleeding from an AVF/AVG.

This study had several limitations. Firstly, this study measured patients’ desires to make decisions, but not the actual decision itself; this may introduce self-reporting or social desirability biases. However, we attempted to mitigate these effects using validated methods including stating the research purpose and assuring confidentiality.(38) Secondly, our patient respondents were predominantly Caucasian, from a single academic Canadian nephrology center, which may limit generalizability of results. Thirdly, this study could not establish if preferences related to Kidney-CAP differed depending on patient characteristics such as race, sex, past kidney transplant, or anticoagulation status. On the other hand, this study was sufficiently powered to make several important conclusions related to the impact the Kidney-CAP had on patient’s choices around kidney transplantation and vascular access. Furthermore, the study established a rational timeline to introduce the Kidney-CAP into the CKD clinic setting. Due to the small sample size, caution should be taken in assuming a causal effect by the Kidney-CAP on desire to proceed with transplantation that we describe in patients receiving hemodialysis.

## CONCLUSIONS

This is the first study evaluating the impact of a bleeding cessation device, for hemodialysis patients with AVF or AVG, on clinical decisions related to vascular access, kidney transplantation, and dialysis modality. The Kidney-CAP was associated with patient interest in both kidney transplantation and creation of AVF. Further study is required to delineate patient decisions within demographic subgroups such as previous kidney transplant, or anticoagulation status.

## Acknowledgements

None

## Author Contributions

Each author contributed to the experimental design. Authors AF and BT completed data collection and analysis. Authors AF and BT completed the first draft of the manuscript, with every author contributing to improvements of subsequent drafts.

## Competing interests

Dr. Ben Thomson is an unpaid medical consultant for Glia Inc, which created the Kidney-CAP. There are no other conflicts of interest.

## Data availability statement

Anonymized dataset is available by contacting the corresponding author.

## Exclusivity statement

There are no related manuscripts under consideration or published elsewhere.

## Funding

None

## Review preferences

No opposition to any specific reviewer.

Yee Gary Ang (Scopus 56364351900), Auron M (Scopus 25026927900), Yuri Battaglia (Scopus 22955424300), Davide Bolignano (Scopus 22133459100), Nasar Alwahaibi (Scopus 35097088600) could all be considered reasonable choices to review.

## APPENDICES

## Appendix 1: Description of the Kidney-CAP Device provided to all study participants, prior to survey initiation

### Description of the Kidney-CAP Device

The **Kidney-CAP** is a new medical device that is designed to help a person who undergoes hemodialysis treatments manage a bleed from a fistula or graft, when that bleed occurs outside the dialysis unit. This is the first device of its kind in North America. A photograph of its use is shown below.

The **Kidney-CAP** attaches to a keychain so that it is always close to or with the patient at all times.

If a patient has bleeding from a fistula or graft, he or she should act **FAST**

**F Find** the Kidney-CAP on your keychain

**A Apply** the Kidney-CAP to bleeding fistula or graft

**S Secure** the Kidney-CAP with tape

**T Telephone** 911

While Kidney Care UK has provided a similarly purposed device to hemodialysis programs, the Canadian design team wanted to evaluate how and when the **Kidney-CAP** should be introduced in the chronic kidney disease pathway. The survey’s results will better inform nephrology programs how and when patients with kidney disease should be introduced to the **Kidney-CAP.**

The **Kidney-CAP** has been designed and produced in a Health Canada approved facility (Glia Inc). We will not be patenting this design because patients everywhere should have access to effective and inexpensive health care. We also provide the **Kidney-CAP** to nephrology programs using a cost recovery model of pricing.

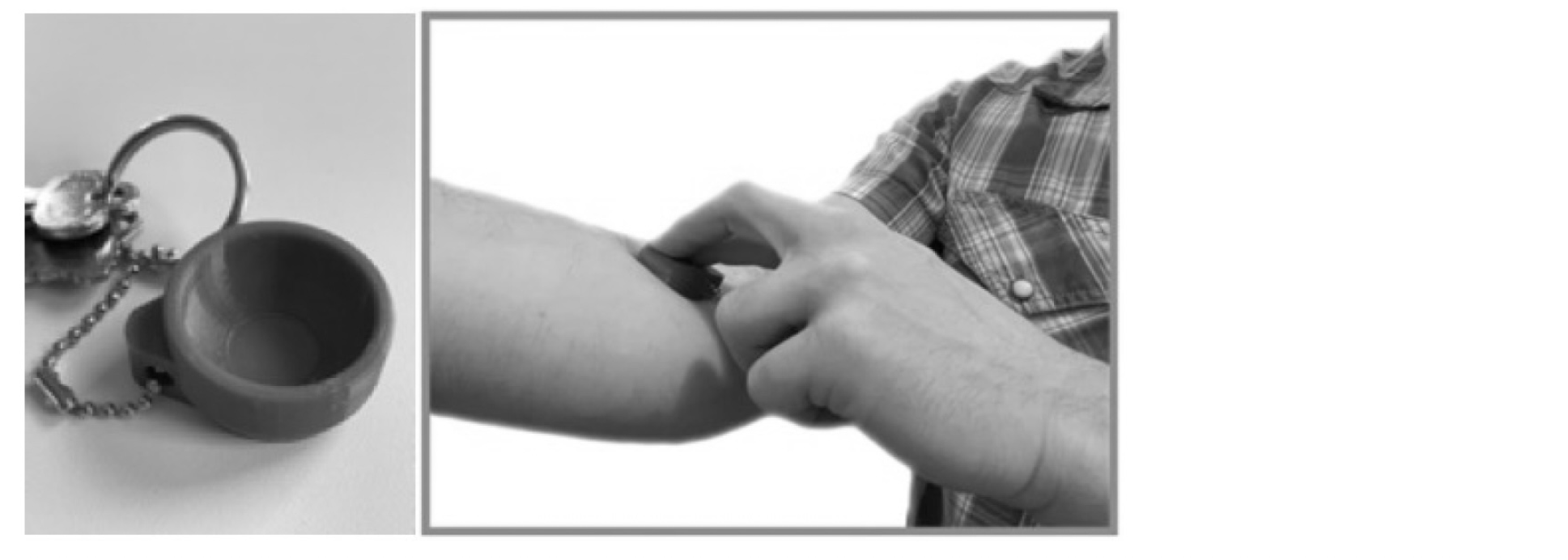

## Notes

### Competing Interest Statement

Dr. Ben Thomson is an unpaid medical consultant for Glia Inc, the company that created the Kidney-CAP device. There are no other conflicts of interests.

### Funding Statement

The author(s) received no specific funding for this work.

## References

1. Wasse H, Gillen DL, Ball AM, Kestenbaum BR, Seliger SL, Sherrard D, et al. Risk factors for upper gastrointestinal bleeding among end-stage renal disease patients. Kidney Int. 2003;64(4):1455–61.

2. Yilnaz U, Unal A, E.E. C, Inci A, Gul S, Demirtas G, et al. Effect of needle bevel position in arteriovenous fistula cannulation and bleeding during hemodialysis. Medicine. 2022;11(3):1311–6.

3. Nursing Recommendations for the Management of Vascular Access in Adult Hemodialysis Patients: 2015 Update. Canadian Association of Nephrology Nurses and Technologists Journal. 2015;25:1–48.

4. Authority BRPHS. Care of Needling Sits Post Hemodialysis for Fistulas and Grafts (Hemostasis). In: Committee BRH, editor. 2020. p. 1–6.

5. Ibeas J, Roca-Tey R, Vallespin J, Moreno T, Monux G, Marti-Monros A, et al. Spanish Clinical Guidelines on Vascular Access for Haemodialysis. Nefrologia. 2017;37 Suppl 1:1–191.

6. Lok CE, Huber TS, Lee T, Shenoy S, Yevzlin AS, Abreo K, et al. KDOQI Clinical Practice Guideline for Vascular Access: 2019 Update. Am J Kidney Dis. 2020;75(4 Suppl 2):S1–S164.

7. Ellingson KD, Palekar RS, Lucero CA, Kurkjian KM, Chai SJ, Schlossberg DS, et al. Vascular access hemorrhages contribute to deaths among hemodialysis patients. Kidney Int. 2012;82(6):686–92.

8. Jose MD, Marshall MR, Read G, Lioufas N, Ling J, Snelling P, et al. Fatal Dialysis Vascular Access Hemorrhage. Am J Kidney Dis. 2017;70(4):570–5.

9. Milosevic E, Forster A, Moist L, Rehman F, Thomson B. Non-surgical interventions to control bleeding from arteriovenous fistulas and grafts inside and outside the hemodialysis unit: a scoping review. Clin Kidney J. 2024;17(5):sfae089.

10. Whitehall C. Put a lid on it: Managing life-threatening bleeds from fistulas and grafts. Kidney Matters. 2019;7:14–6.

11. Karatzas E. ‘Stop the bleed’ and save lives with something as simple as a milk bottle lid. Riotact: Local voices Stronger Communities. 2022 November 1, 2022.

12. Casey JR, Hanson CS, Winkelmayer WC, Craig JC, Palmer S, Strippoli GF, et al. Patients’ perspectives on hemodialysis vascular access: a systematic review of qualitative studies. Am J Kidney Dis. 2014;64(6):937–53.

13. Duncanson E, Le Leu RK, Shanahan L, Macauley L, Bennett PN, Weichula R, et al. The prevalence and evidence-based management of needle fear in adults with chronic disease: A scoping review. PLoS One. 2021;16(6):e0253048.

14. Kosa SD, Bhola C, Lok CE. Hemodialysis patients’ satisfaction and perspectives on complications associated with vascular access related interventions: are we listening? J Vasc Access. 2016;17(4):313–9.

15. McLaughlin K, Manns B, Mortis G, Hons R, Taub K. Why patients with ESRD do not select self-care dialysis as a treatment option. Am J Kidney Dis. 2003;41(2):380–5.

16. Androga LA, Amundson RH, Hickson LJ, Thorsteinsdottir B, Garovic VD, Manohar S, et al. Telehealth versus face-to-face visits: A comprehensive outpatient perspective-based cohort study of patients with kidney disease. PLoS One. 2022;17(3):e0265073.

17. Collister D, Herrington G, Delgado L, Whitlock R, Tennankore K, Tangri N, et al. Patient views regarding cannabis use in chronic kidney disease and kidney failure: a survey study. Nephrol Dial Transplant. 2023;38(4):922–31.

18. Tennankore KK, McCullough K, Bieber B, Cho Y, Johnson DW, Kanjanabuch T, et al. Prevalence and Outcomes of Chronic Kidney Disease-Associated Pruritus: International Results from Peritoneal Dialysis Outcomes and Practice Patterns Study. Clin J Am Soc Nephrol. 2024;19(12):1622–34.

19. Wulczyn KE, Rhee EP, Myint L, Kalim S, Shafi T. Incidence and Risk Factors for Pruritus in Patients with Nondialysis CKD. Clin J Am Soc Nephrol. 2023;18(2):193–203.

20. Acharya D, Scory TD, Shommu N, Donald M, Harrison TG, Murray JS, et al. Nephrologist’s Perceptions of Risk of Severe Chronic Kidney Disease and Outpatient Follow-up After Hospitalization With AKI: Multinational Randomized Survey Study. Can J Kidney Health Dis. 2025;12:20543581251336548.

21. Brimble KS, Mehrotra R, Tonelli M, Hawley CM, Castledine C, McDonald SP, et al. Estimated GFR reporting influences recommendations for dialysis initiation. J Am Soc Nephrol. 2013;24(11):1737–42.

22. Sharma A, Minh Duc NT, Luu Lam Thang T, Nam NH, Ng SJ, Abbas KS, et al. A Consensus-Based Checklist for Reporting of Survey Studies (CROSS). J Gen Intern Med. 2021;36(10):3179–87.

23. Jones EL, Shakespeare K, McLaughlin L, Noyes J. Understanding people’s decisions when choosing or declining a kidney transplant: a qualitative evidence synthesis. BMJ Open. 2023;13(8):e071348.

24. Grams ME, Brunskill NJ, Ballew SH, Sang Y, Coresh J, Matsushita K, et al. The Kidney Failure Risk Equation: Evaluation of Novel Input Variables including eGFR Estimated Using the CKD-EPI 2021 Equation in 59 Cohorts. J Am Soc Nephrol. 2023;34(3):482–94.

25. Multi-Care Kidney Clinic Best Practices. Ontario Renal Network; 2022.

26. Nizic-Kos T, Ponikvar A, Buturovic-Ponikvar J. Reasons for refusing kidney transplantation among chronic dialysis patients. Ther Apher Dial. 2013;17(4):419–24.

27. Griva K, Seow PS, Seow TY, Goh ZS, Choo JCJ, Foo M, et al. Patient-Related Barriers to Timely Dialysis Access Preparation: A Qualitative Study of the Perspectives of Patients, Family Members, and Health Care Providers. Kidney Med. 2020;2(1):29–41.

28. Murray MA, Thomas A, Wald R, Marticorena R, Donnelly S, Jeffs L. Are you SURE about your vascular access? Exploring factors influencing vascular access decisions with chronic hemodialysis patients and their nurses. CANNT J. 2016;26(2):21–8.

29. Woo K, Pieters H. The patient experience of hemodialysis vascular access decision-making. J Vasc Access. 2021;22(6):911–9.

30. De Siqueira J, Jones A, Waduud M, Troxler M, Stocken D, Scott DJA. Systematic review of interventions to increase the use of arteriovenous fistulae and grafts in incident haemodialysis patients. J Vasc Access. 2022;23(5):832–8.

31. Shamasneh AO, Atieh AS, Gharaibeh KA, Hamadah A. Perceived barriers and attitudes toward arteriovenous fistula creation and use in hemodialysis patients in Palestine. Ren Fail. 2020;42(1):343–9.

32. Xi W, Harwood L, Diamant MJ, Brown JB, Gallo K, Sontrop JM, et al. Patient attitudes towards the arteriovenous fistula: a qualitative study on vascular access decision making. Nephrol Dial Transplant. 2011;26(10):3302–8.

33. Arenas MD, Cazar R, Cordon A, Mendez A, Acuna M, Furaz K, et al. Is it possible to reach the catheter target proposed by the guidelines? Reasons for catheter use in prevalent hemodialysis patients. Nefrologia (Engl Ed). 2024;44(5):700–8.

34. Israni AK, Halpern SD, McFadden C, Israni RK, Wasserstein A, Kobrin S, et al. Willingness of dialysis patients to participate in a randomized controlled trial of daily dialysis. Kidney Int. 2004;65(3):990–8.

35. Agoritsas T, Perneger TV. Patient-reported conformity of informed consent procedures and participation in clinical research. QJM. 2011;104(2):151–9.

36. Locock L, Smith L. Personal benefit, or benefiting others? Deciding whether to take part in clinical trials. Clin Trials. 2011;8(1):85–93.

37. Welton AJ, Vickers MR, Cooper JA, Meade TW, Marteau TM. Is recruitment more difficult with a placebo arm in randomised controlled trials? A quasirandomised, interview based study. BMJ. 1999;318(7191):1114–7.

38. Ried L, Eckerd S, Kaufmann L. Social desirabiilty bias in PSM surveys and behavioral experiments: Considerations for design development and data collection. Journal of Purchasing and Supply Management 2022;28(1):100743.

